# Perceived Health Benefits of Using Non-Electrical Water Filters under ‘Clean India Mission’ Programme in a Village of Bhopal

**DOI:** 10.1101/2020.11.26.20238451

**Authors:** Sindhuprava Rana, Vikas Dhiman, Nalok Banerjee, Anil Prakash, Rajnarayan R Tiwari

## Abstract

In India, the access to clean and safe drinking water to rural population is very limited, hence the Government of India has initiated multiple schemes to tackle the problem of huge health burden due to poor quality of water in rural areas. Under ‘Clean India Mission’ cost-effective, non-electric water filters were distributed (n=144 households) in Barkheda Bondar village of Bhopal district in Central India and after one year of usage, a questionnaire based door-to-door survey was conducted to assess the usage and perceived health benefits of water filters by the users. The study showed that the acceptance of water filter among rural population was about 82% and 69.4% of the population reported reduction in the frequency of various water-related diseases. The present study shows that the use of non-electric water filters on mass basis in rural India is efficacious for providing safe drinking water.

## Introduction

In India, the access to clean drinking water is a fundamental right under Article 21 of the Constitution of India and ensuring availability and sustainable management of water and sanitation for all is one of the Sustainable Development Goals (SDGs) adopted by the United Nations (Anand and Roy, 2016). As per the report of National Rural Drinking Water Programme (India), only 44% of the rural households in India had access to safe drinking water in 2017 (https://cag.gov.in/). It has been estimated that about 37.7 million Indians are affected by waterborne diseases, 1.5 million children die of diarrhoea and 73 million working days are lost annually leading to an economic burden of $600 million a year (Mudur, 2003). Hence the health burden due to poor quality of water and its supply in rural India is enormous (Mudur, 2003). In a recent study regarding water treatment practices in rural India showed that 68.6% of the study participants do not treat water before usage resulting in various waterborne diseases (Ravindra et al., 2019). Multiple studies have shown that the use of cheap, cost-effective water filtration techniques in rural areas reduces the contamination of water and waterborne diseases (Rao et al., 2003; Freeman et. al, 2012; Francis et al., 2016). The Government of India has thus initiated multiple schemes to tackle the problem of huge health burden due to poor quality of water in rural areas.

One of such initiatives ‘Swachh Bharat Abhiyan’ or ‘Clean India Mission’ was launched on October 2, 2014 with an aim to achieve a clean and Open Defecation Free (ODF) India by 2nd October 2019. One of the objectives of this mission is to encourage the use of cost effective technologies for safe and sustainable sanitation including drinking water. Contributing towards ‘Swachh Bharat Abhiyan’, this institute distributed non-electric water filters in a village of Bhopal district in Central India after assessing the felt needs of the villagers with the assumption that use of such cost-effective water filters in rural households will ensure clean and sustainable source of clean drinking water in rural areas. In the present study, we present the usage and perceived health benefits of the water filters among rural population after one year of distribution of water filters in a village.

## Methods

### Study design

This is a cross-sectional community based survey, which was carried out in Barkheda Bondar village (23.30°N and 77.29°E) under Phanda block of Bhopal district, Madhya Pradesh. According to 2011 census of India, it had 435 households with total population of 2005 (Males: 1019, females: 986). The literacy rate of the population (excluding children ≤ 6 years) is 64.61%. Majority of the working population of this village are agricultural laborers (48.4%) (http://www.censusindia.gov.in/). Under Swachh Bharat Abhiyan, we carried out various activities including cleanliness drive, providing dustbins for wet and dry garbage collection, awareness campaign etc. in this village. It was seen that many households in the village were using water from the open dug-wells for drinking purposes without any treatment. Further, it was noticed that the villagers had no knowledge about the need for purification of water before drinking and cooking. On finding the poor quality of water being used for drinking in the village, the need to distribute water filters was felt and thereafter, water filters based on Ultra-filtration/Electro Adsorption Technology (EAT) with multi-stage purification process was selected for distribution based on their non-dependence on electricity and cost-effectiveness.

In March 2018, one water filter per household was distributed, covering 144 households, based on the information collected from the head of the village. Prior to the distribution, a cleanliness campaign was organized to demonstrate the fitting, usage and cleaning of water filters to the users. After one year of the distribution of water filters, a survey was carried out covering all 144 beneficiary households in March 2019 to assess the extent of usage and perceived health benefits of the water filters by the household members. All the households, to whom the water filters were distributed, were included in the survey. The informed signed consent was taken from each house hold surveyed. The survey was conducted by a team of doctors using a semi-structured questionnaire in Hindi containing 22 questions pertaining to socio-demographic information and water filter usage (Table 1). The questionnaire was adapted from the previous published questionnaires (Denslow et al., 2010; Ordinioha B, 2011). Hindi is the vernacular language spoken by the villagers and the survey team; hence there was no language barrier between the survey team and the villagers. The use of the water filters was physically verified by the investigators in the households and the response of the users were obtained from the head/senior member of the family. The information was recorded in the pre-designed digital excel spreadsheet. The data was tabulated and descriptive analysis was done using SPSS software *ver* 21.

## Results

### Population

After one year of distribution, 124 households could be contacted (coverage: 86.1%) covering a population of 854 (males: 452, females: 402). Twenty households were found to be locked/ shifted to another place, so responses could not be elicited from these households. The facility of toilet was present in 119 households (96.0%), while two-third of the houses were kachha built (n=84, 67.7%).

### Drinking water storage practices

A total of 74 (59.7%) and 44 (35.5%) households were using bore-well and open dug-well as source of drinking water respectively. Only a few houses were using tap water (2 households, 1.6%) or from other sources (4 households, 3.2%). Majority of the households stored drinking water in plastic containers or *kuppi* (n=79, 63.7%) or metal/steel pot (n=26, 21.0%). Other common modes of storage of water were water filter (n=10, 8.1%) and earthen pots (n=7, 5.6%). Nearly 50% households (n=63) were not using any treatment before storing the drinking water. Among those using any kind of treatment, sieving (n=50, 40.3%) was the most common method. Only a few households used boiling (n=3, 2.4%) as water treatment option.

### Usage of water filter

The water filters were physically verified in use in 102 (82.3%) households. Rest of the households either did not use (n=22, 17.7%) or abandoned after using for sometime (n=2, 1.6%). The reasons cited for non-usage of water filters were non-palatable taste of water, breakage, or gifting to relatives among others.

### Perceived health benefits

The frequency of water-borne diseases was felt to be reduced after using water filters in 86 households (69.4%), remained static in 18 households (14.5%) while no response could be elicited from 20 households (16.1%). Most of the households (n=41, 33.1%) reported reduction in the frequency of multiple diseases like diarrhoea, fever, stomach pain, skin diseases etc. The reduction in the frequency of diseases like upper respiratory infections (n=12, 9.7%), diarrhoea (n=7, 5.6%), fever (n=6, 4.8%) and itching (n=2, 1.6%) were also reported. Sixty three percent households reported that they recommended others (relatives, neighbors, colleagues, etc.) to start using water filters for water treatment while 26 households (21.0%) responded in negative.

## Discussion

Usage of water filtration is known to reduce signs and symptoms like stomach pain, flatulence, diarrhoea, fatigue etc. (Rao et al., 2003; Freeman et. al, 2012). It has been reported that after consuming water from community installed water filters in a village in West Bengal, India reported 7.1-21.5% decrease in various water-related diseases (Majumdar, 2011). The present study adds to the existing knowledge about the perception of health benefits as perceived by the rural population after using non-electric water filters distributed under ‘Clean India Mission’ campaign, in a village in Central India.

The population in rural India is not accustomed to usage of filters for drinking water. Among the first time users, the acceptance of water filters was found reasonably high (n=102/144, 82.3%) and 84% (n=86/102) of the users perceived one or the other health benefits after using water filters, which shows that providing non-electric, inexpensive water filters may be a useful tool to reduce the burden of water-borne diseases in the rural settings in India. Eleven households (8.9%) reported no benefits of the usage of water though the exact reason for the same could not be evaluated. It is striking that over 50% households in the present study were not using any kind of water treatment methods before storage, which shows the lack of awareness about safe drinking water practices in the community. Similar situations were seen in the urban/semi-urban areas of Bhopal city (Banerjee et al., 2020) and also in the rural areas of Chandigarh, India (Ravinder et al., 2019). Though the villagers commonly use sieving method through a cloth as water purification technique but the use of water filters certainly has an edge over the conventional methods of sieving in terms of health benefits.

Majority of the households use plastic containers to store drinking water in rural areas (64%), which is similar to urban areas (Sruthy and Ramasamy, 2017). The good old Indian tradition of using pitchers/earthen pots for water storage that not only cools the water (Huage et al., 1994) but also cleans the water was found in use in only 6% of the households. It has been shown in the present study that about two-third of the houses in Barkheda Bondar are *kachha* but 96% of the households were having toilet facilities indicating an increase of awareness of cleanliness and sanitation among rural population in India. The source of drinking water for majority of the village is groundwater, 35.5% of the households use open wells for fetching water that may be a potential source of contamination of water. The recent initiative by the Government of India to provide “Piped Water for All by 2024” will likely reduce dependence of villagers on open wells for drinking water.

The limitation of the present study is small sample size and the data on health problems were collected as they were perceived by the villagers. Nevertheless, the study suggests that promoting the use of non-electric water filters in rural areas may effectively improve the quality of drinking water, thus, bringing down the burden of water-borne diseases.

## Conclusion

The present study shows that the use of non-electric water filters on mass basis in rural India is efficacious for providing safe drinking water. Such practices may be implemented in rural areas of Africa and Asia where there is poor quality of water and high burden of waterborne diseases.

## Data Availability

All the survey data is being stored in the study institute

## Author contribution

SR proposed the conception idea for the study. SR and VD screened and extracted the data. SR analyzed the data and drafted the manuscript. NB provided the necessary feedback. VD, AP and RRT edited the manuscript.

## Acknowledgements

We acknowledge the contributions of the members of the Swachh Bharat Committee of ICMR-NIREH, Bhopal. Last but not the least; we are grateful to the subjects of Barkheda Bondar village for allowing us to carry out this study.

## References

Anand A, Roy N. Transitioning toward Sustainable Development Goals: The Role of Household Environment in Influencing Child Health in Sub-Saharan Africa and South Asia Using Recent Demographic Health Surveys. Front Public Health 2016;4;87.

Banerjee N, Banerjee A, Sabde Y, Tiwari RR, Prakash A, NIREH Epidemiology Research Group. Morbidity profile of communities in Bhopal city (India) vis-a-vis distance of residence from Union Carbide India Limited plant and drinking water usage pattern. J Postgrad Med 2020;66:73–80.

Denslow SA, Edwards J, Horney J, Peña R, Wurzelmann D, Morgan D. Improvements to water purification and sanitation infrastructure may reduce the diarrheal burden in a marginalized and flood prone population in remote Nicaragua. BMC Int Health Hum Rights. 2010 Dec 8;10:30. doi: 10.1186/1472-698X-10-30.

Francis MR, Sarkar R, Roy S, Jaffar S, Mohan VR, Kang G, et al. Effectiveness of Membrane Filtration to Improve Drinking Water: A Quasi-Experimental Study from Rural Southern India. Am J Trop Med Hyg 2016;95;1192–200.

Freeman MC, Trinies V, Boisson S, Mak G, Clasen T. Promoting household water treatment through women’s self help groups in Rural India: assessing impact on drinking water quality and equity. PLoS ONE 2012;7;e44068.

Hauge S, Osterberg R, Bjorvatn K, Selvig KA. Defluoridation of drinking water with pottery: effect of firing temperature. Scand J Dent Res 1994;102;329–33.

Majumdar KK. Health impact of supplying safe drinking water containing fluoride below permissible level on flourosis patients in a fluoride-endemic rural area of West Bengal. Indian J Public Health 2011;55;303–8.

Mudur G. India’s burden of waterborne diseases is underestimated. BMJ 2003; 14;326;1284.

Ordinioha B. A Survey of the Community Water Supply of some rural Riverine Communities in the Niger Delta region, Nigeria: Health implications and literature search for suitable interventions. Niger Med J. 2011;52:13–18.

Ravindra K, Mor S, Pinnaka VL. Water uses, treatment, and sanitation practices in rural areas of Chandigarh and its relation with waterborne diseases. Environ Sci Pollut Res Int 2019;26;19512–22.

Rao RR, Reddy RC, Rao KGR, Kelkar PS. Assessment of slow sand filtration system for rural water supply schemes--a case study. Indian J Environ Health 2003;45;59–64.

Sruthy S, Ramasamy EV. Microplastic pollution in Vembanad Lake, Kerala, India: The first report of microplastics in lake and estuarine sediments in India. Environ Pollut 2017;222;315–22.

